# Systematic review of precision subclassification of type 2 diabetes

**DOI:** 10.1101/2023.04.19.23288577

**Authors:** Shivani Misra, Robert Wagner, Bige Ozkan, Martin Schön, Magdalena Sevilla-Gonzalez, Katsiaryna Prystupa, Caroline C. Wang, Raymond J. Kreienkamp, Sara J. Cromer, Mary R. Rooney, Daisy Duan, Anne Cathrine Baun Thuesen, Amelia S. Wallace, Aaron Leong, Aaron J. Deutsch, Mette K. Andersen, Liana K. Billings, Robert H Eckel, Wayne Huey-Herng Sheu, Torben Hansen, Norbert Stefan, Mark O. Goodarzi, Debashree Ray, Elizabeth Selvin, Jose C. Florez, ADA/EASD PMDI, James B. Meigs, Miriam S. Udler

**Author notes:** Corresponding address &. These authors contributed equally as first author (Shivani Misra, Robert Wagner). ^#^Joint senior authors (James B. Meigs, Miriam S. Udler). ^&^See acknowledgements block (ADA/EASD PMDI).

## Abstract

Heterogeneity in type 2 diabetes presentation, progression and treatment has the potential for precision medicine interventions that can enhance care and outcomes for affected individuals. We undertook a systematic review to ascertain whether strategies to subclassify type 2 diabetes are associated with improved clinical outcomes, show reproducibility and have high quality evidence. We reviewed publications that deployed ‘simple subclassification’ using clinical features, biomarkers, imaging or other routinely available parameters or ‘complex subclassification’ approaches that used machine learning and/or genomic data. We found that simple stratification approaches, for example, stratification based on age, body mass index or lipid profiles, had been widely used, but no strategy had been replicated and many lacked association with meaningful outcomes. Complex stratification using clustering of simple clinical data with and without genetic data did show reproducible subtypes of diabetes that had been associated with outcomes such as cardiovascular disease and/or mortality. Both approaches require a higher grade of evidence but support the premise that type 2 diabetes can be subclassified into meaningful groups. More studies are needed to test these subclassifications in more diverse ancestries and prove that they are amenable to interventions.

## Introduction

Type 2 diabetes is a global health problem posing a significant burden on healthcare systems. By 2045, ∼11% of the world’s population (783 million individuals) is projected to have a diagnosis of type 2 diabetes^1^. The prevention, better treatment and improved outcomes for people with type 2 diabetes is a major health priority.

The diagnosis of type 2 diabetes is based on haemoglobin (HbA1c), fasting plasma glucose, or oral glucose tolerance test coupled with the absence of clinical features that may indicate an alternative diabetes subtype (such as type 1 diabetes, monogenic, pancreatic or medication-induced)^2^. A diagnostic label of type 2 diabetes is therefore either arrived at through exclusion of other subtypes or in most cases, is the default diagnosis. Following a diagnosis of type 2 diabetes, most national and international guidelines have advocated the same treatment algorithms irrespective of individual patient characteristics and treatment choice has been based on features such as cost, side effects, or desire of weight loss, that are not necessarily related to the underlying pathophysiology. More recently international guidelines have suggested differential glucose-lowering therapies on the basis of higher body mass index (BMI) (favouring use of glucagon-like peptide analogue, GLP-1) and presence or absence of cardiovascular and/or renal disease and/or heart failure (GLP-1 and/or sodium-glucose co-transporter 2, SGLT-2 inhibitors)^3^.

Significant heterogeneity in the clinical presentation, individual characteristics and progression of patients with type 2 diabetes, is recognised at the clinical interface, including differences in degree of obesity, age, dyslipidaemia, presence of metabolic syndrome and’or body fat distribution^4–6^. Approaches such as clustering methods, to categorise this heterogeneity have been deployed and shown differences in progression to complications or need for insulin treatment. These approaches consider characteristics at diagnosis^7^ or combined clinical information with genetic data to demonstrate disease heterogeneity^8, 9^. There is now growing recognition that type 2 diabetes heterogeneity may reflect variability in the underlying pathophysiology, environmental contributors and the genetic risk of affected individuals; that the risk factors and mechanisms leading to the development of type 2 diabetes may differ significantly from one individual to another and this could impact treatments. .

The goal to individualise care with more precise treatment in order to achieve better outcomes drives the impetus to capture and categorise this heterogeneity.g. Such a goal has been realised in part for monogenic diabetes, where treatments can be tailored to genetic subtype to deliver precision diabetes care achieving better outcomes than standard care^10^. Given the complex pathophysiology of type 2 diabetes and its variable polygenic risk, applying precision medicine approaches to the care of affected individuals is more challenging but may still be achievable. Critical to this endeavour is a better understanding of specific subtypes in order to achieve precise subclassification of type 2 diabetes.

Studies of type 2 diabetes subtypes have spanned a long time period and included a multitude of approaches based on the presence, absence or a specific threshold of one or more simple clinical features or biomarkers and, recently, more sophisticated methods that deploy machine learning (ML) and/or genetic or other ‘omics data. ‘Simpler’ approaches have potential merit in being easily deployed across all resource settings, whilst complex approaches may have greater benefit in drilling deeper beyond outward clinical features to decipher and classify heterogeneity. The breadth and scope of the evidence in favour of type 2 diabetes subclassification has not to date been thoroughly examined.

In this systematic review for the *Precision Medicine in Diabetes Initiative* (PMDI) we aimed to provide a comprehensive and critical assessment of the evidence to date for type 2 diabetes subclassification using (i) simple approaches based on clinical features, biomarkers, imaging, or other routinely available parameters, and (ii) complex subclassification approaches that incorporate ML, genomic data, and clinical data. We aimed to identify areas where further research is needed to develop more accurate, cost-effective, and clinically relevant strategies for the subclassification of type 2 diabetes, ultimately leading to improved patient outcomes and a more efficient allocation of healthcare resources.

The PMDI was established in 2018 by the American Diabetes Association (ADA) in partnership with the European Association for the Study of Diabetes (EASD). The ADA/EASD PMDI includes global thought leaders in precision diabetes medicine who are working to address the burgeoning need for better diabetes prevention and care through precision medicine^11^. This Systematic Review is written on behalf of the ADA/EASD PMDI as part of a comprehensive evidence evaluation in support of the 2^nd^ International Consensus Report on Precision Diabetes Medicine [crossref Tobias et al, Nat Med].

## Methods

This systematic review was written and conducted in accordance with our pre-established protocol (PROSPERO ID CRD42022310539) and reported using the Preferred Reporting Items for Systematic Reviews and Meta-Analyses Statement (PRISMA)^12^. We systematically reviewed papers to address two research questions devised by an expert working group; 1) What are the main subtypes of type 2 diabetes defined using simple clinical criteria and/or routinely available laboratory tests (simple approaches), and 2) What subphenotypes of type 2 diabetes can be reproducibly identified using ML and/or genomics approaches in different populations (complex approaches)? The quality of each paper was reported, and the aggregate of data evaluated using the GRADE system^13^.

### Study Eligibility Criteria

We included English-language studies of all designs that undertook analysis of populations with prevalent or new-onset type 2 diabetes and attempted in some way to stratify or subgroup type 2 diabetes. We used broad terms to identify stratification studies and all approaches to stratification (the exposure) were included (see supplementary). We excluded studies examining risk for the development of type 2 diabetes, use of glycemia alone to stratify, or studies of stratification in types of diabetes other than type 2 diabetes, and review articles or case reports.

Simple approaches were defined as follows:

1) The exposure used inputs from any of the following

a) a routine blood or urine biomarker that was widely available in most clinic settings
b) a blood or urine biomarker that might not be routinely available now but could have the potential to become easily accessible
c) any routinely available imaging modality
d) any physiological assessment that could be undertaken in an outpatient setting
e) results from routinely available dynamic tests
2) The stratification approach was

a) a cut-off or categorisation based on one or more of the above
b) if an index, ratio, trend or other analysis was undertaken, it could be calculated without complex mathematics
3) Outcomes

a) All outcomes were accepted

Complex approaches were defined as follows:

1) The exposure used

a) Any of the inputs for the simple approach outlined above and/or
b) Any form of genetic data
2) The stratification approach either

a) Deployed ML approaches or
b) Used other complex statistical approaches for stratification
3) Outcomes

a) All outcomes were accepted

### Literature Search & Selection Strategy

PUBMED and EMBASE databases were searched from their inception to May 2022 for relevant articles using a strategy devised by expert health sciences librarians (see appendix for search terms used). We undertook two separate searches for each systematic review question. Each abstract and subsequently, full text paper, were screened by two independent team members for eligibility. In addition to the initial exclusion criteria, at the full-text review stage, we further excluded studies where the *only* stratification modality was a measure of glycaemic control, where exposures were not clearly defined and/or if the data on outcomes of the stratification were not available in results or supplementary material. Conflicts were resolved by group consensus and/or a third independent reviewer.

### Data Extraction

Data were manually extracted from each full-text paper by individual team members and cross-checked by an independent team member at the data synthesis stage. We extracted relevant data on study design (observational or clinical trial), analysis design (cross-sectional or prospective), study population characteristics, stratification method (exposure), outcomes assessed, and study quality assessment. For population characteristics we extracted data on whether the type 2 diabetes population was new-onset or prevalent, the sample size, the ethnicity and gender of those studied, the duration of diabetes (for cross-sectional analysis) and duration of follow-up (for longitudinal follow-up). For exposure, we extracted the number of subgroups identified, the approach to stratification and subgroups identified. For outcomes, we documented the type of outcome studied and the findings according to stratified subgroup.

### Data Synthesis

Following full-text data extraction, we undertook a qualitative analysis of exposures (measures used to stratify individuals with type 2 diabetes) for each systematic review question. For simple approaches to subclassification, we extracted the details of stratification criteria in each paper (see **supplementary table 1**), then agreed on the category of exposure this fell into (e.g. blood / urine test, imaging, age etc). After data extraction, these exposures were further refined into subcategories based on common emerging themes (e.g. use of pancreatic autoantibodies, BMI categories, measures of beta-cell function, use of lipid profiles, etc.). For complex approaches, the exposure was both the input clinical and/or genetic data used and the ML approach to analysis (e.g. k-means, hierarchical clustering, latent-class analysis, etc.), deployed. In both simple and complex systematic reviews, outcomes studied were heterogeneous. These outcomes were later broadly categorised where possible. Due to the variability in exposures and outcomes, it was not possible to undertake meta-analysis of any outcomes, however main conclusions of the relationship between exposures and outcomes in similar studies were evaluated. All coding, categorisation and thematic analysis was undertaken and agreed by at least three members of the research team.

### Quality Assessment

The GRADE system was used to assess the quality of the studies extracted^13^. As well as assessing whether exposures and outcomes were clearly defined, valid and reliable, at least two members of the research team assessed whether confounders were appropriately accounted and adjusted for. Recognising the majority of studies were observational or case control, assessors used these two assessments, along with broad assessment of study limitations, inconsistency of results, imprecision and reporting bias to assign GRADE certainty rating as very low, low, moderate and high^14^.

## Results

### Search and screening (for Q1 and Q2)

The first question examined simple stratification approaches using clinical variables that may explain diabetes heterogeneity. A total of 6097 studies met the inclusion criteria and were screened (see **figure 1a**). 183 studies were included for full text data review of which 132 studies were subsequently excluded. The most common reasons for exclusion at the full-text review stage, were studies in populations without prevalent or incident type 2 diabetes, a study design that used ML approaches or stratification approaches that used HbA1c or diabetes medications. In total 51 studies underwent full-text data extraction.

**Figure 1.**
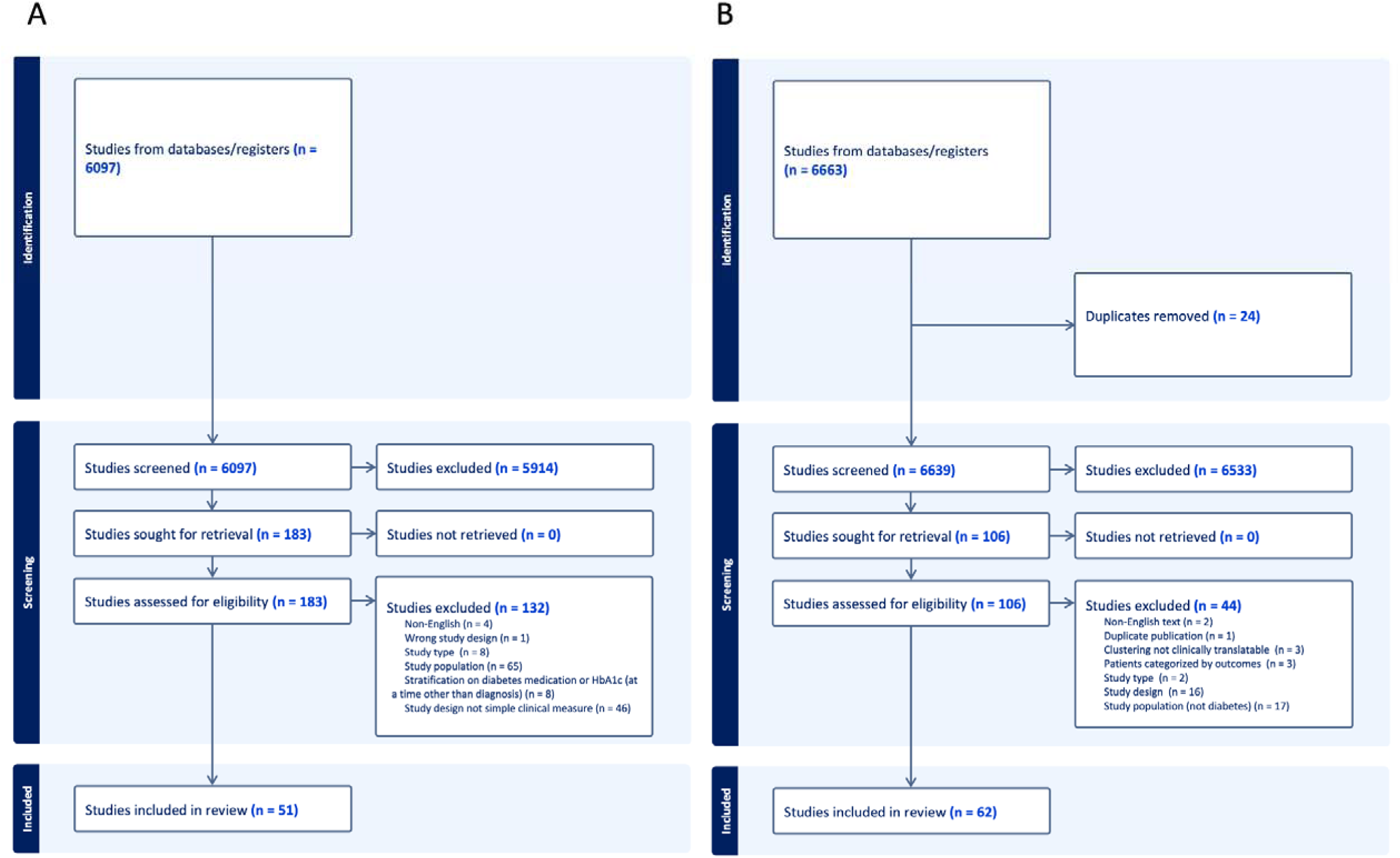
A, B: Systematic review attrition diagram. Left panel (A) for simple approaches to subclassification and right panel (B), complex approaches.

The second question aimed to identify papers with complex approaches, mostly ML based strategies, to identify subgroups of type 2 diabetes (see **figure 1b**). A total of 6639 studies were screened, of which 106 were found eligible for full-text review. The most common reasons for exclusion were study populations not comprising participants with type 2 diabetes or classification approaches not using ML. After full-text screening, 62 papers were included in the systematic review.

#### Use of simple approaches to subclassify type 2 diabetes

### Description of extracted studies

The 51 studies using simple type 2 diabetes subclassification approaches incorporated 1,751,350 participants with prevalent or new-onset type 2 diabetes. 39% (20/51) of studies included participants of white European ancestry, 43% (22/51) incorporated exclusively participants from non-white European ancestries and 17% (9/51) included mixed ancestry groups (**supplementary Table 1)**. The majority of the studies (78%, 40/51) were conducted in populations with prevalent type 2 diabetes, and 22% (11/51) in new-onset type 2 diabetes. Approximately half the studies had a prospective design (25/51), the remaining half had a cross-sectional (26/51) design. For longitudinal studies, the follow-up period ranged from <1 year to 22 years.

Studies included wide range of exposures (see **Figure 2** for summary) based on routine clinical measurements with standard cut-offs or groupings; continuous assessment of individual routine clinic based measurements e.g. variability over time; composite stratification incorporating 2 or more tiers of criteria e.g. cut-offs or groupings with one or more biomarkers or anthropometric measurements; stratification using alternate modalities including non-routine but clinically available biomarkers or imaging or clinical scores or responses to oral glucose tolerance tests (OGTT). The associations of stratified exposure characteristics were investigated with various outcomes: 1) measures of glycaemia, 2) clinical characteristics, 3) measures of diabetes progression such as time-to-insulin treatment or development of microvascular complications and 4) cardiovascular outcomes and/or mortality.

**Figure 2:**
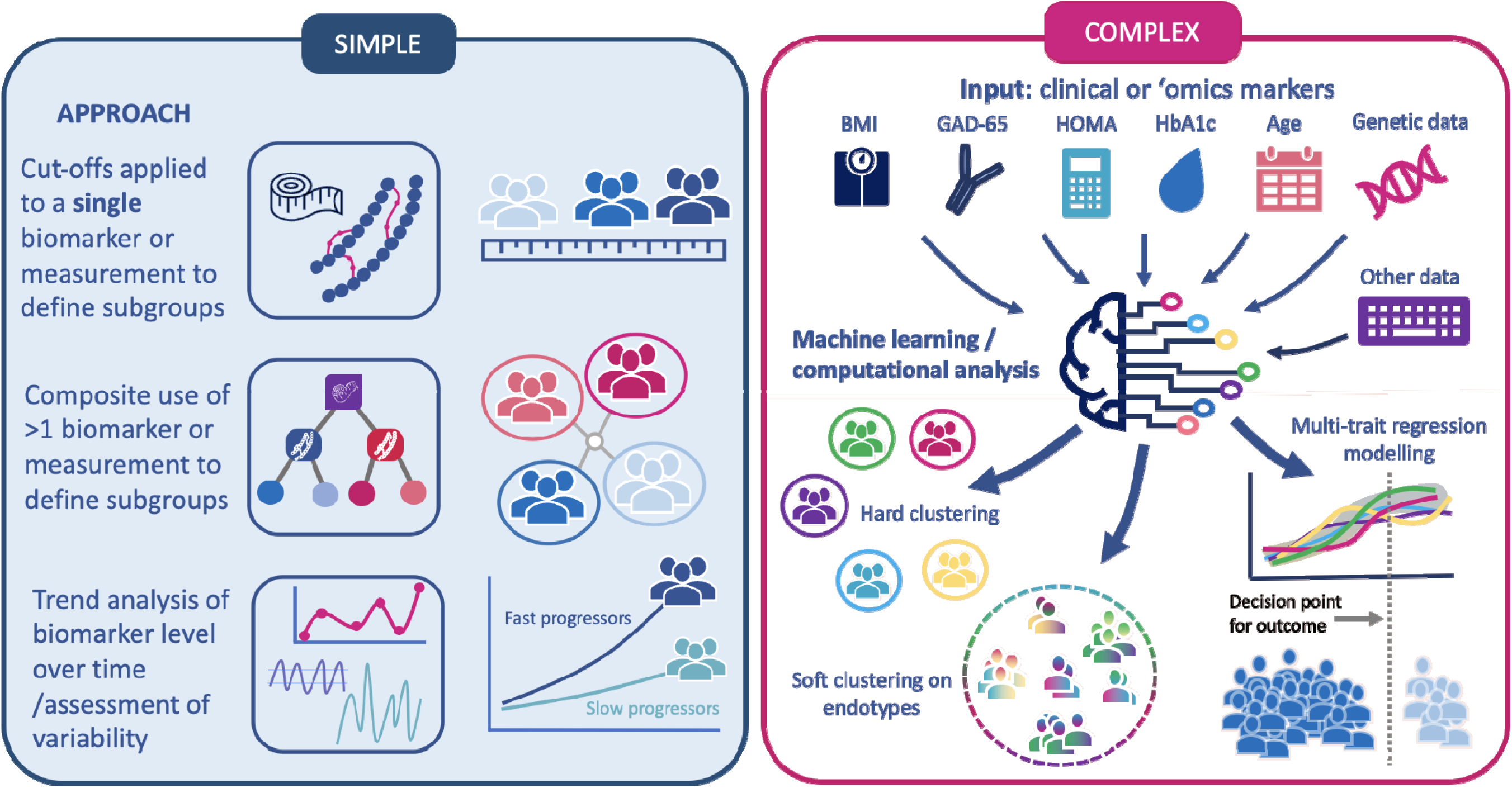
Schematic overview of approaches used to subclassify type 2 diabetes. Left panel depicting ‘simple approaches’ to type 2 diabetes subclassification and right panel ‘complex approaches’. HbA1c, glycated haemoglobin; BMI, body mass index; GAD-65, glutamic acid decarboxylase-65 antibodies.

### Description of categorised subgroups

Simple approaches to classification included use of lipid profiles (n=8), BMI (n=6), pancreatic beta-cell related measures (n=6), pancreatic auto-antibodies (n=6), age at diagnosis (n=2), OGTT data (n=4), cardiovascular measures (n=3), other biomarkers in urine or blood and alternative approaches (n=5) (see **Table 2** for summary).

Different categories of triglycerides, low-density lipoprotein (LDL) cholesterol, high-density lipoprotein (HDL) cholesterol, atherogenic small dense lipoproteins with and without features of metabolic syndrome were used to stratify type 2 diabetes in 8 studies. Cardiovascular disease outcomes were assessed in 3/8 of the studies^15–17^ which showed that a more atherogenic metric of the specific lipid exposure (e.g. higher LDL cholesterol) was associated with a greater frequency of CVD outcomes. Other outcomes included pulsewave velocity^18^ or clinical characteristics; age, BMI, presence of metabolic syndrome in specific subgroups.

The six studies assessing pancreatic autoantibodies focused on GAD-65 levels. Studies used positive versus negative status or high versus low titre, and one study sub-stratified by age. Outcomes included time-to-insulin treatment ^19, 20^, associations with other clinical characteristics such as lipid profiles, BMI and blood pressure^21–23^ and measures of beta-cell function. There was no consistency in study design and most were observational with low to moderate evidence grade; two studies showed that GAD-65 positivity was associated with faster-time-to insulin treatment^19, 20^.

Patients with type 2 diabetes were stratified according to their BMI in six studies, either by itself (n=5) or in combination with HbA1c. The number of BMI categories varied between two and six in the identified studies. The association between BMI and glycaemic outcomes (change in HbA1c from baseline) were assessed in four studies either as primary or secondary outcomes^5, 24, 25^. We graded the quality of evidence as very low to moderate, and there was no consistency of effect observed. In one secondary analysis of a randomised control trial, higher BMI at baseline was associated with faster progression to renal outcomes, however this was not replicated in any other study^26^.

Age at diagnosis of type 2 diabetes was assessed as a stratification tool in two studies; younger age (mean age 33 years) was associated with higher rates of peripheral retinopathy in an observational study with 12 months follow-up versus older aged (mean 50 years)^4^. In a second study, people aged 60-75 versus those >75 years had a high risk of CVD and mortality when stratified by cholesterol levels^6^.

OGTT was an exposure in four studies. The specific stratification approach applied to OGTT profiles was different in each study and based on cut-offs of fasting glucose levels, glucose gradients after stimulation and responses to different drug treatments. Outcomes included clamp-derived insulin sensitivity and differences in the shape of glucose profiles between youth and adults^27^.

Beta-cell measures were assessed in six studies including C-peptide levels and homeostasis model assessment-2 indices for beta-cell function (HOMA2-B) or insulin resistance (HOMA2-IR). C-peptide was defined using variable cut-offs. Outcomes included clinical phenotype data, response to medication, and microvascular or macrovascular complications.

Other exposure variables included less routine biomarkers, pulse wave velocity, ketosis/ketoacidosis and other disease indices, however single studies prevented grouping of these exposures - all data summarised in **Table** 1.

**Table 1.**
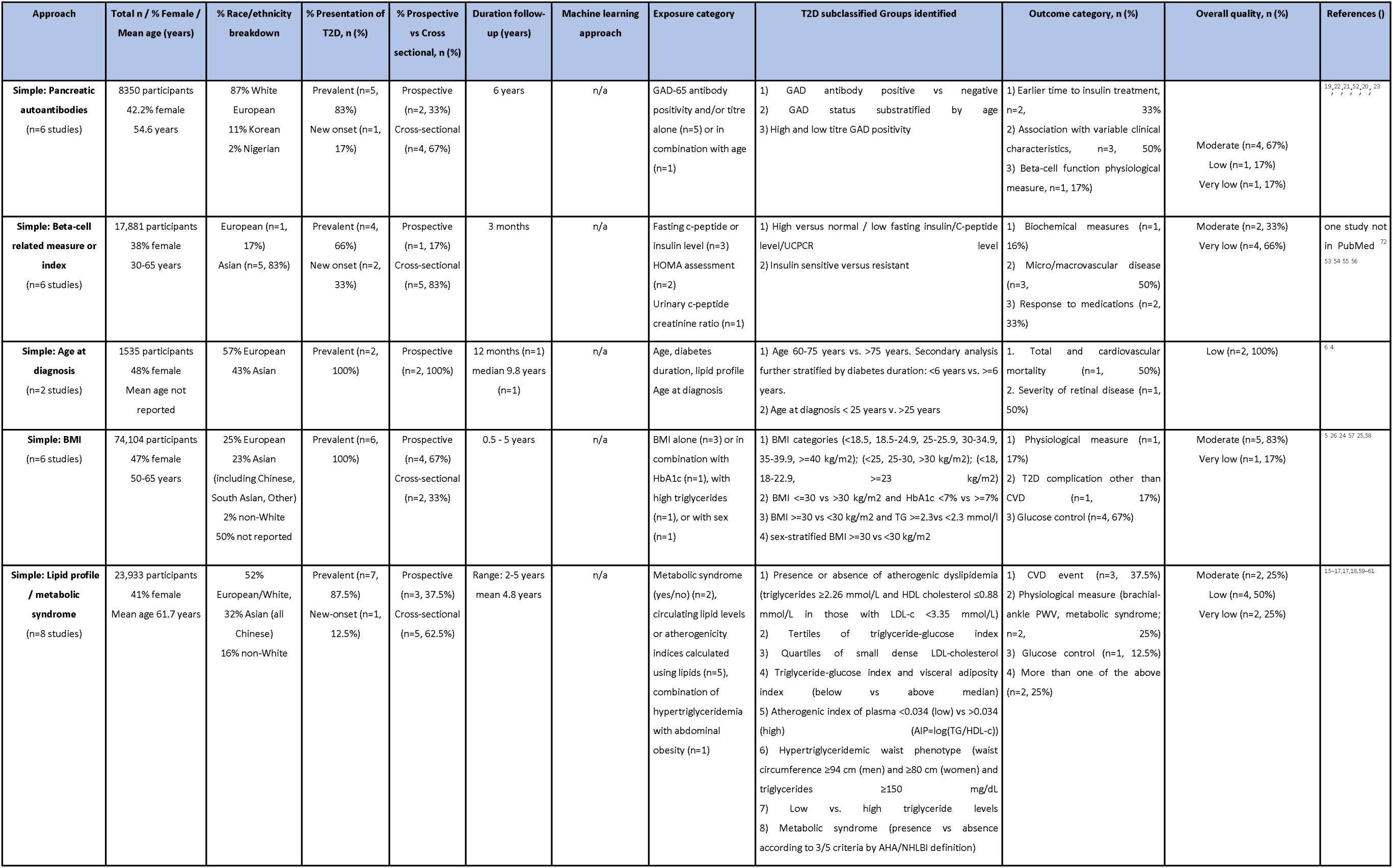
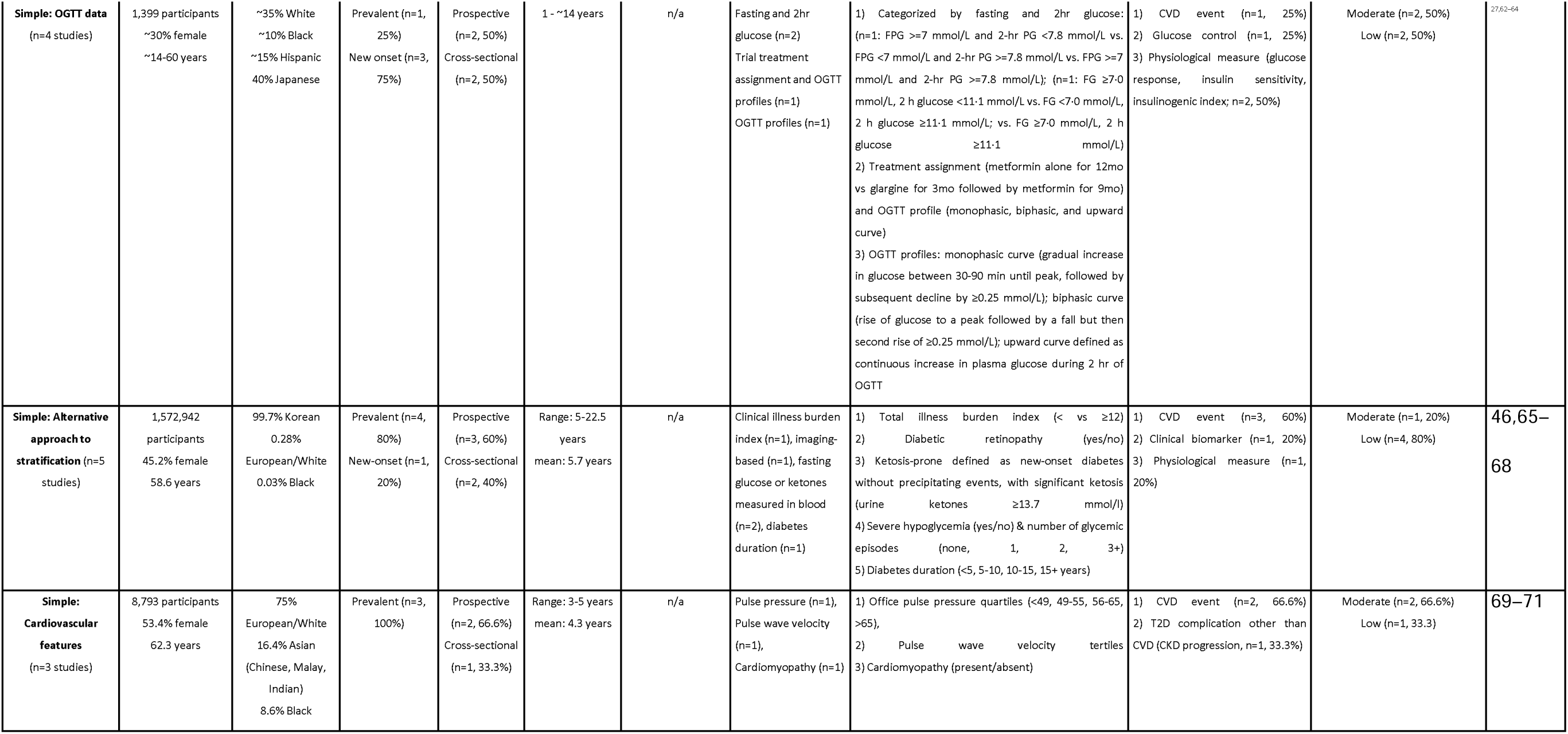
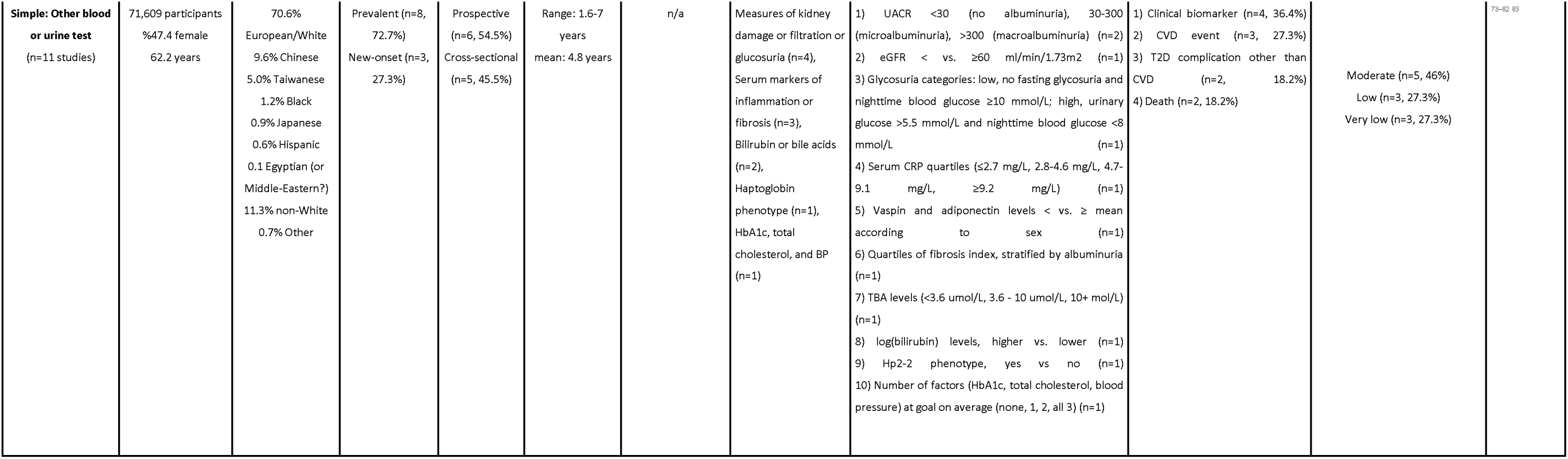

Of the 51 studies assessed, 55% were graded as very low, or low GRADE certainty, 45% had moderate certainty and none achieved high certainty.

### Use of complex approaches to subclassify type 2 diabetes

#### Description of extracted studies

There were 62 studies of complex/ML approaches to type 2 diabetes subclassification in a total of 793,291 participants (**Table 2**). Over half of the studies included non-European ancestry in relevant proportions (>20%). Only ∼30% (19 out of 62) of the studies analysed participants with new-onset diabetes. Mean diabetes duration ranged from recent onset (within 1 year) to over 36 years. Most data were from observational studies (46 out of 62), with some post-hoc analyses of clinical trials (10), survey data (4) and mixed study types (2). Half of the studies had prospective design (31 out of 62) with a mean follow-up duration ranging from 1 year to 11.6 years. K-means clustering was the most commonly applied ML approach (30/62). Eight studies used established centroids^7^ to assign participants to clusters. Two studies decomposed combinations of genetic variants and their association with clinical and laboratory phenotypes into genotype-phenotype clusters by using Bayesian non-negative matrix factorization.

**Table 2.**
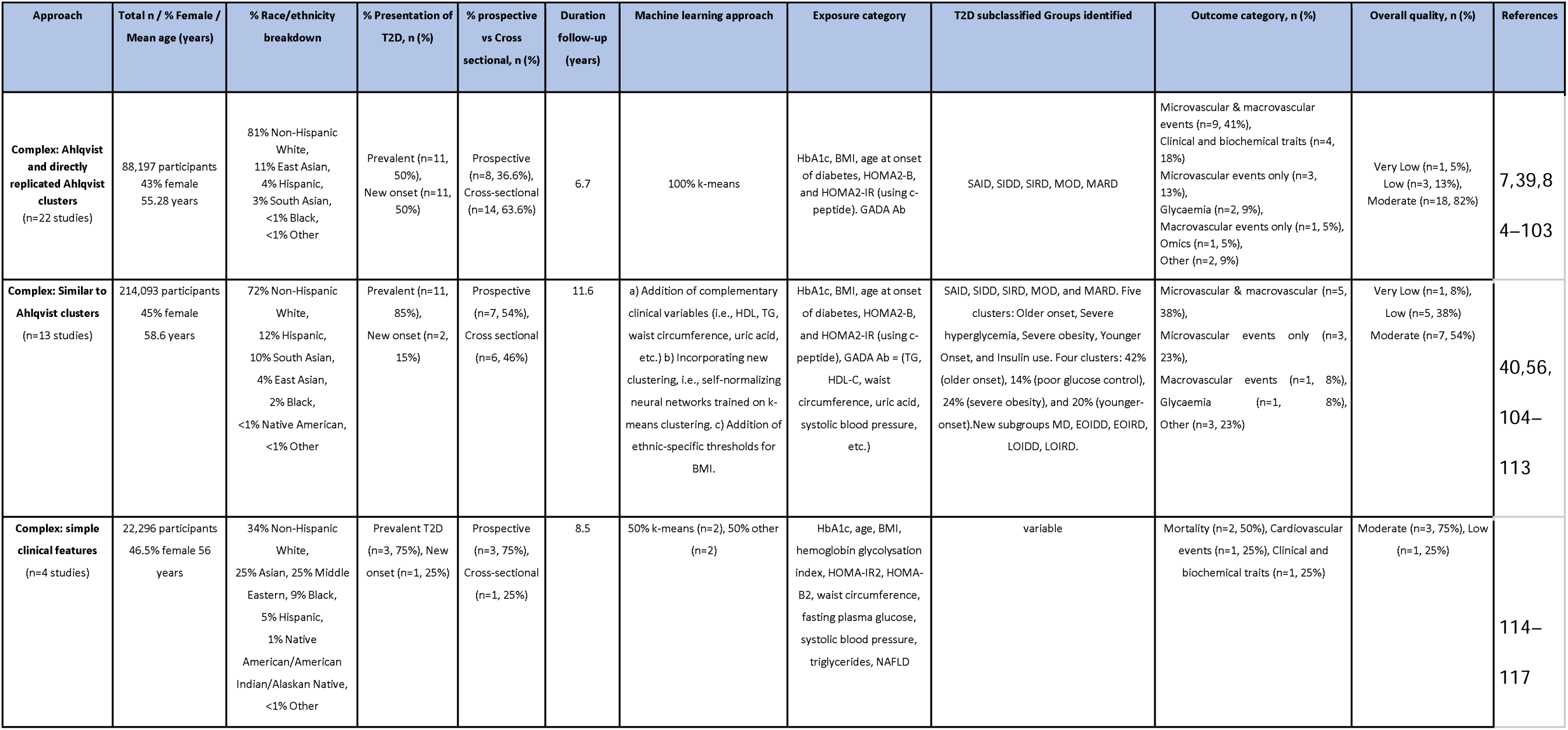
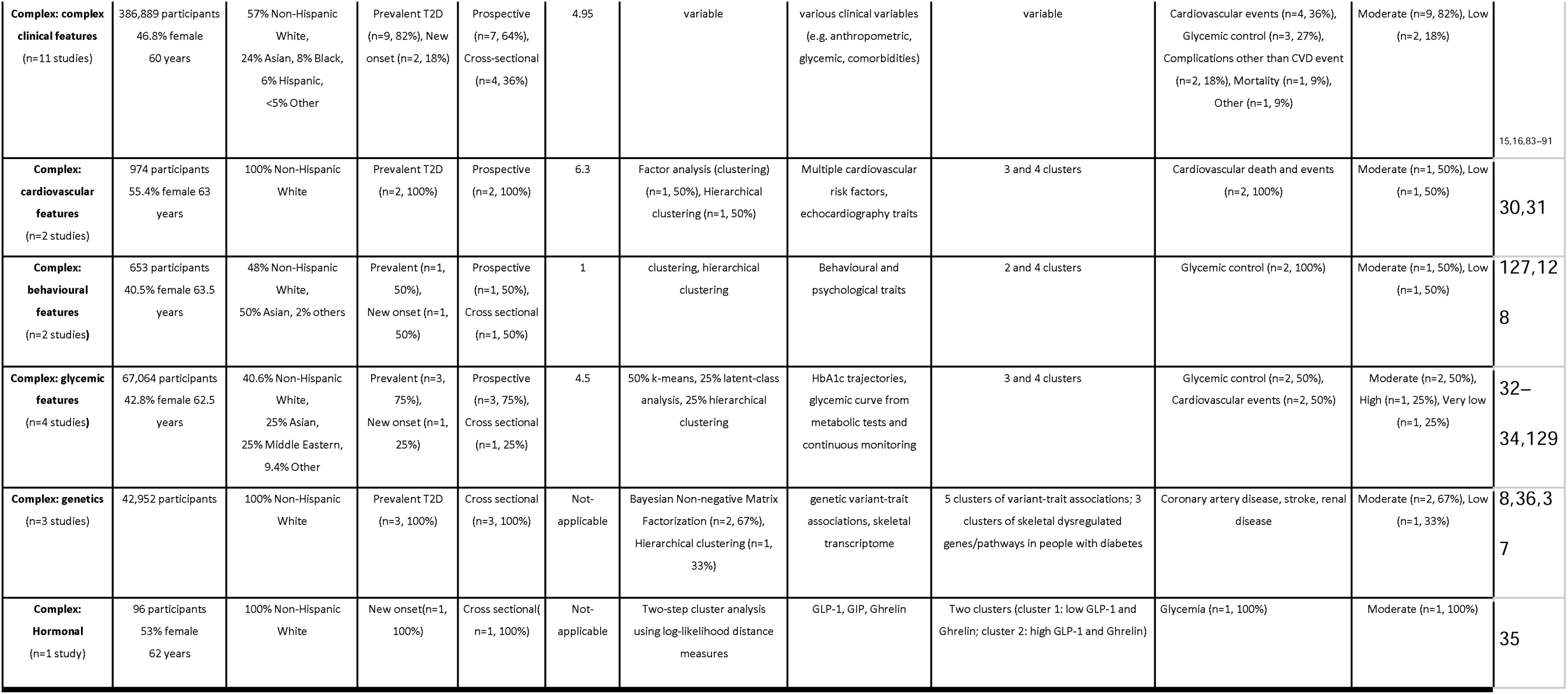

#### Description of the categorised subgroups

Following up on the seminal work by Ahlqvist *et al.* ^7^, multiple studies used the variables age of diabetes onset, HbA1c, BMI, HOMA2-B, HOMA2-IR and GAD-65 antibody for diabetes subclassification (**Table 2**). The majority of these studies employed C-peptide-based homeostasis model assessment indices (HOMA, or its updated variant, HOMA2 (which require measurement of fasting insulin and glucose), as surrogates for insulin resistance (HOMA2-IR) and insulin secretion (HOMA2-B). Twenty-two studies replicated identification of the five diabetes subtypes described in Ahlqvist *et al.*^7^ in different contexts and populations: severe insulin-deficient diabetes (SIDD), severe insulin resistant diabetes (SIRD), mild obesity-related diabetes (MOD) and mild age-related diabetes (MARD) with or without the GAD-positive severe autoimmune diabetes (SAID) subtype. Associations of these subtypes with clinical outcomes including glycaemic traits, microvascular and macrovascular outcomes and death replicated in 12 studies (**Table 3**).

**Table 3.** Association of the Ahlqvist-clusters with outcomes from 22 reviewed studies using consistent cluster assignment methods

|  | Glycemic outcomes | Microvascular outcomes | Macrovascular outcomes | Other outcomes | Death |
| --- | --- | --- | --- | --- | --- |
| **SIDD** | Insulin requirement<sup>7,92</sup> | Retinopathy<sup>7,92,97-99</sup><br>(N=3160)<br>Nephropathy<sup>92,98</sup><br>(N=1074) | Lower extremity arterial disease<sup>98</sup> (N=223) | Distal symmetric polyneuropathy, Cardiac autonomic neuropathy<sup>85</sup> (N=28), Erectile dysfunction<sup>88</sup> (N=4) |  |
| **SIRD** | Glycemic benefit with thiazolidinedione therapy<sup>39</sup> (N=800) | Diabetic kidney disease<sup>7,85,93,95,98,102</sup><br>(N=2686) | MACE (confounded by age and sex<sup>7</sup>, N=1373) | Prevalence of NAFLD<sup>85</sup> (N=121), Erectile dysfunction<sup>88</sup> (N=7), Diabetic peripheral neuropathy<sup>98</sup> (N=225) |  |
| **MOD** |  |  |  | Neuropathy<sup>92</sup> (N=1258) |  |
| **MARD** | Glycemic benefit with sulfonylurea therapy<sup>39</sup><br>(N=1361) | Retinopathy<sup>95</sup> (N=1487) | MACE<sup>7,92</sup> (N=3513) |  | CVD mortality<sup>97</sup> (N=56) |

Thirteen additional papers used variations of the original set of variables from Ahlqvist *et al.* ^7^ by substituting HOMA with C-peptide, adding lipid traits e.g. HDL, or approximating the clusters from different / simplified variable sets by applying advanced statistical learning approaches such as self-normalizing neural networks. These approaches identified some type 2 diabetes subgroups resembling the clusters from Ahlqvist *et al.* and also novel subgroups related to the additional variables (**Figure 3**). Several of the novel subgroups were associated with clinical outcomes, however, these findings were not replicated in other studies (**Table 2**).

**Figure 3:**
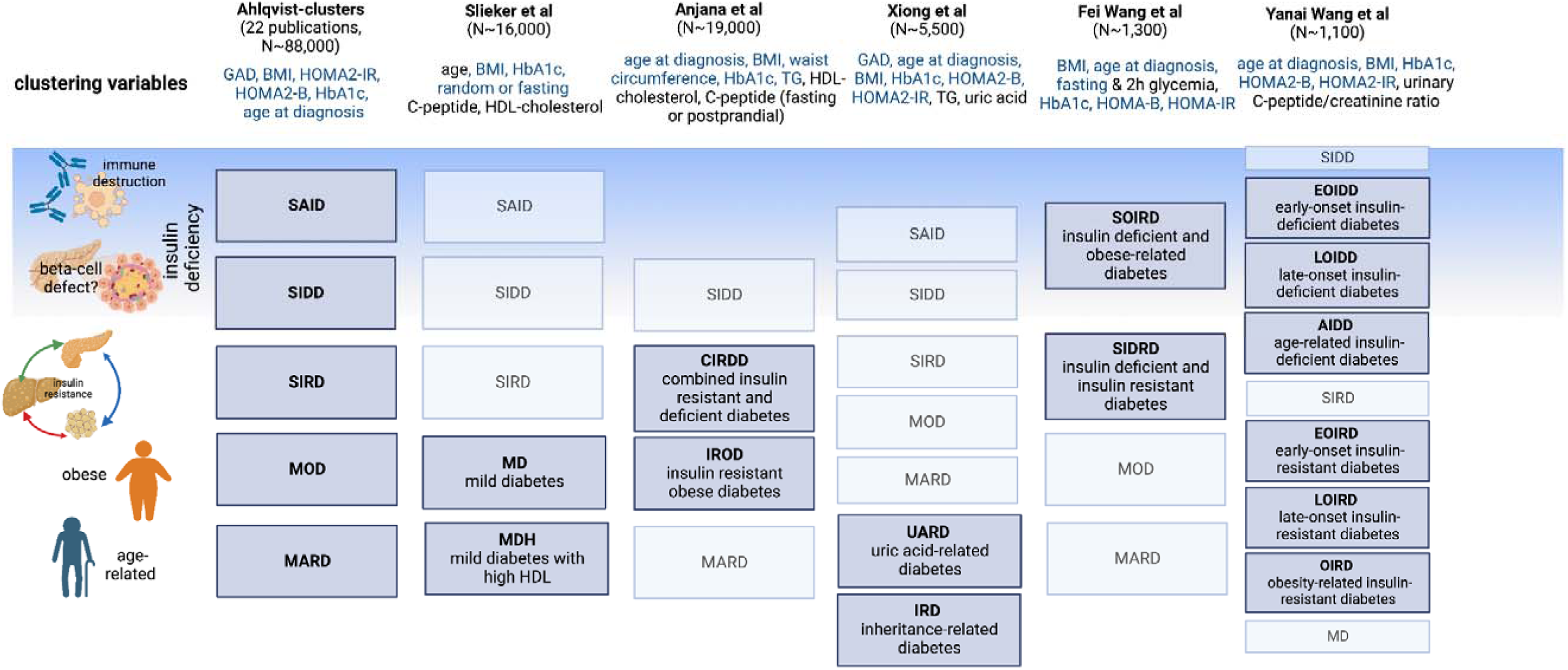
Main characteristics of diabetes clusters derived using different clustering variables, compared to original ‘Alhqvist’ clusters. Clustering variables denoted in blue are consistent across the different studies, those in black are unique to the particular study outlined. A greyed out box indicates that the indicated diabetes cluster was replicated from the Ahlqvist study, a dark blue box indicates a new diabetes cluster.

Additional papers (n=27) assessed various sets of phenotypic inputs for ML approaches. These studies, grouped here into five categories of inputs, identified many subtypes of diabetes and associations with clinical outcomes; however, they all lacked replication (**Table 2**). Four papers applied complex ML methods to a set of less than ten clinical variables such as systolic blood pressure, waist circumference, BMI, fasting plasma glucose, and age at diabetes diagnosis, and resulting subgroups were associated with outcomes such as mortality. Eleven studies used a larger set of more than ten clinical features as inputs for classification, including data from electronic health records^28, 29^, and identified subgroups with associated clinical outcomes, including risk of cardiovascular disease. Two other studies specifically employed cardiovascular traits including ECG^30^ and echocardiographic^31^ for ML algorithm inputs, and each identified subgroups with different associations with risk of cardiovascular disease. Finally, four studies involved inputs of change of glycaemic variables (HbA1c trajectories, glycaemia during a mixed meal test, continuous glucose monitoring features), ^32–34^ one study focused on fasting GLP-1, GIP and ghrelin levels^35^, and two studies focused on behavioural traits such as novelty seeking, harm avoidance, and hospital anxiety and depression scale.

Two sets of papers utilised genomic data to identify diabetes subtypes, either in the form of inherited common genetic variation ^8, 36^ or gene expression data from muscle biopsies^37^ (**Table 2**). The former approach started with genetic variants associated with type 2 diabetes, and clustered their genotype-phenotype associations to identify subsets of variants predicted to act in shared mechanistic processes. Using these sets of genetic variants, “process-specific” or “partitioned polygenic scores” were constructed in individuals with type 2 diabetes, yielding clusters for insulin resistance and insulin secretion that were associated with differences in clinical features and prevalence of metabolic outcomes, with replication across multiple cohorts. Of note, however, despite reaching statistical significance, the associations observed between genetic subtypes (defined as individuals with the top 10% of process-specific polygenic risk) and others with type 2 diabetes were too small for clinical utility at the individual level.

Overall, half of the studies had cross-sectional designs, and the other half involved prospective follow-up(**Table 2**). Around 70% of the studies had moderate evidence certainty.

## Discussion

### Summary of Findings

This systematic review comprehensively and qualitatively analysed two broad approaches to the subclassification of type 2 diabetes to identify clinically meaningful subtypes and advance precision diagnostics. Simple approaches to subclassification were common and included diabetes-related urine and blood biomarkers, anthropometric measures, clinical data such as age at diagnosis, surrogate beta-cell metrics derived from C-peptide or insulin along with other less intuitive exposures such as non-diabetes related biomarkers (e.g. bilirubin levels) or pulse wave velocity. Approaches to subclassification were diverse; some studies used dichotomous separation of a a continuous variable based on a clinical cut-point; in others, two or more criteria were applied in a composite approach and analysis of variability or trends of continuous data were also used e.g. change in eGFR over time.

Although many exposures were tested, the study designs, specific cut-offs used and outcomes were heterogenous and no studies were of high quality GRADE certainty. It was not possible to assess reproducibility of type 2 diabetes subtypes using data from studies that deployed simple approaches, however, some studies discovered biologically plausible type 2 diabetes subgroups. For example, in our qualitative analysis, subclassifications derived using BMI, beta-cell function, lipid profiles and age appeared to be associated with a number of meaningful outcomes which could be helpful in clinical practice. These potential subclassifications need to be pursued in better designed studies (see section on additional supporting literature). Other evidence not specifically included in our systematic review questions do support the role of simple variables in stratifying diabetes, for example, younger age at diagnosis is reproducibly associated with worse cardiorenal outcomes in a number of epidemiological analysis^38^.

Machine learning approaches yielded some reproducible subtypes of type 2 diabetes using a variety of clinical and genetic variables. The best-replicated subtypes were the five clusters first described in Ahlqvist *et al.*^7^, which were replicated in 22 studies including approximately 88,000 individuals of diverse ancestry. Additionally, there was replication of genetic subtypes of type 2 diabetes from Udler *et al.*^8^ with clinical features seen in multiple cohorts across almost 454,000 individuals^36^. However, these associations involved small absolute effects which challenges clinical utility for individual patient management, and these studies were restricted to European ancestry individuals. While there was replication of the clusters from Ahlqvist *et al.* across studies, the generated clusters appeared to be dependent on the characteristics of the underlying populations, especially factors such as distribution of ancestry, age, duration of diabetes, anthropometric trait variability as in BMI, and the variety of variable terms included in learning models. Nevertheless, at least some of the resulting subtypes appeared to be robust to differences in specific ML method, input variables, and populations (**Figure 3**).

Many of the input variables for the complex ML subtyping approaches were also used in studies involving simple approaches to subclassification, recapitulating the biological plausibility of specific clustering variables in defining type 2 diabetes subtypes. One study directly compared a simple clinical approach to the clustering approach from Ahlqvist *et al.* and found that simple single clinical measures analysed in a quantitative (rather than categorical) framework could better predict relevant clinical outcomes, such as incidence of chronic kidney disease and glycemic response to medications^39^. Thus, further research is needed to determine whether assigning a patient to one of the clusters from Ahlqvist *et al.* offers additional clinical benefit beyond evaluation of simple clinical measures and also beyond current standard of care. For example, high quality randomised controlled trial evidence is needed to prove that for differential treatment and/or clinical care pathways knowledge of a patient’s cluster membership makes a material difference.

### Study Quality

No studies included in our systematic review had above moderate certainty of evidence. Strengths of the included studies were their large sample sizes, the diversity of variables considered, and inclusion of both prevalent and new-onset cases of type 2 diabetes. However, the varied study designs and lack of replicationblimits the ability to draw clear conclusions about the most effective approaches to subclassification. Most variables used for subclassification capture momentary metabolic states, and cluster assignment can change over time, therefore lacking stability and sufficient predictive accuracy^40, 41^. Additionally, most studies focused on European-ancestry populations, and the clinical value of these approaches may vary across different ancestries. It was positive to see that some ancestries have representations such as East Asians, however replication in Black, south Asian and Hispanic populations is definitely required. This is particularly important, as four out of five people with type 2 diabetes hail from marginalised groups or live in low or middle income countries. Finally, most studies were retrospective, and there are no data involving subtype-stratified clinical trials or “real world” implementation of approaches. Future precision diagnostic interventions should address, not widen inequalities.

### Additional supporting literature

Since our literature search was conducted, new publications have advanced our understanding of type 2 diabetes subclassification.

Two recent studies applied ML approaches to stratify diabetes heterogeneity, both considering continuous approaches rather than with discrete clusters^42,43^. Nair, et al. (2022) used a non-linear transformation and visualisation of nine variables onto a tree-like structure^43^ and with replication in two large datasets^42^. This approach linked underlying disease heterogeneity to risk of complications, e.g. those at risk of cardiovascular disease had a different phenotype to those with microvascular complications and to drug response, and also demonstrated associations of gradients across the tree with the genetic process-specific scores from Udler et al.8. Wesolowska-Andersen et al. performed soft-clustering from 32 clinical variables which yielded 4 diabetes archetypes comprising ∼⅓ of the study population. The remaining study population was deemed as mixed-phenotype. This study has not been replicated^42^.

Additionally, one of the first clinical trials to assess precision medicine approaches for diabetes management was published. The Trimaster Study tested dichotomised BMI and eGFR strata in a three-period crossover trial using three pharmacologic interventions with the primary hypothesis being stratum-specific differences in HbA1c^44^. Participants with obesity (BMI > 30 kg/m2) showed a glycemic benefit on pioglitazone vs. sitagliptin and participants with lower eGFR (between 60 and 90 ml/min/1.73m2) responded with lower HbA1c to sitagliptin as compared to canagliflozin. In a secondary analysis, drug-choice corresponding to patient preferences yielded lower glycemia than a random allocation, suggesting that “listening to patients” is critical in informing therapeutic decisions^45^. Ramifications of this study are limited by the non-comparable pharmacologic doses used, and the primary focus on glycaemia which may not be indicative of long-term therapeutic success and/or prevention of complications. Yet, these studies have generated higher quality evidence linking type 2 diabetes heterogeneity to treatment and disease outcomes, and it remains to be seen if these can be replicated in other ancestries and translated into ‘usable products’ for healthcare professionals.

It is worth noting that ketosis-prone type 2 diabetes, an established type 2 diabetes subtype, was not captured adequately in our systematic review; only one study evaluated ketosis-prone type 2 diabetes as an exposure^46^. Study designs for ketosis-prone type 2 diabetes are usually analyses of cohorts with diabetic ketoacidosis at presentation, rather than as an exposure in people with type 2 diabetes. Since our search was designed to identify studies stratifying type 2 diabetes, these distinct cohort studies were not captured. Like many other ‘simple’ criteria for classification, the clinical characteristics of people with diabetic ketoacidosis at presentation of type 2 diabetes have been studied, but prospective follow-up analyses are few^47^ and even then, moving forwards beyond phenotype to differences in outcomes or treatment, have not been realised.

Age-at-diagnosis as a simple approach to stratification also did not feature strongly in our search results. The body of literature that outlines higher risk of microvascular or macrovascular complications in early-onset type 2 diabetes has focussed on comparing people with type 2 diabetes to those without diabetes in different age groups^38,48^ or studied cohorts of early-onset cases in isolation^49^ and thus would not have been captured in our search strategy. Recent epidemiological publications have not directly compared outcomes between early and late age onset strata^50,51^ showcasing higher risks of cardiorenal outcomes with early-age at onset, but these are retrospective analyses of health record databases and are potentially confounded by age-related risk of complications and duration of diabetes. To move forward, prospective studies stratifying different interventions (e.g. tighter treatment targets or better cardiovascular risk reduction) by age-at-diagnosis are needed, to prove clinical benefit of age-related subclassication.

### Findings in context

Our analysis has shown that simple features have not been evaluated to a high enough standard to subclassify type 2 diabetes into subtypes more precisely and reproducibly. Perhaps this is not surprising as many studies were not necessarily conducted with ‘precision diagnosis’ in mind, but rather as studiess of clinical phenotypes spanning a time period before the concept of ‘precision diagnosis’ evolved. These studies have ‘set the scene’, but the field needs more robust evidence. It is important to note that many of the simple clinical criteria studied, do have other bodies of evidence supporting associations with outcomes, such as age-at-diagnosis as outlined above.

In contrast, ‘complex’ methods (which emerged with a precision diagnosis label very much in mind) have shown better reproducibility, have been linked to a variety of meaningful clinical outcomes more consistently, and more recently have been able to demonstrate differential treatment responses related to stratification.

What does this mean for a precision medicine approach to type 2 diabetes diagnosis? Ideally these strategies should be deployed at diagnosis of type 2 diabetes on the basis of clinical characteristics and/or ‘omics, such that people in different subgroups or with ‘different flavours’ of type 2 diabetes could be treated differently. One key question is whether such efforts would actually improve clinical outcomes compared to the current standard of care and clinical usefulness and cost-effectiveness of such approaches would need to be evaluated.

For most subclassification approaches described, there is a need for replication in independent datasets, assessment in more diverse populations, in people with both new-onset and prevalent diabetes, and investigation using prospective data, ideally in the form of randomised clinical trials. Additionally, high-quality evidence regarding the therapeutic utility of diabetes subphenotypes could be obtained from randomised controlled studies examining cluster-specific treatment effects, which are yet to be carried out. Ideally, subphenotyping should lead to stratification of complication risks, and such approaches should incorporate the prevention of diabetes complications as endpoints. However, conducting these studies may be challenging due to the necessity for extensive follow-ups, large sample sizes, and significant resource requirements. There is a pressing need for innovative strategies to generate high-quality evidence on treatment options tailored to specific diabetes subtypes in diverse populations. These data will be critical to determine generalizability of findings and amenability for clinical translation.

### Clinical Applicability

The current evidence supports that there are robust subtypes of type 2 diabetes and that these subtypes are associated with clinical outcomes. However, the low to moderate quality of existing studies and the need for replication in ancestry-diverse studies make it difficult to identify a strongly evidence-based, universally applicable approach.

The most clinically valuable methods are likely to be those that are easy and inexpensive to implement. For more complex approaches, decision support tools will need to be developed and assessed for feasibility and utility. Although the evidence supporting complex approaches has, effectively, leap-frogged the evidence in favour of more simplified approaches, there is still likely to be a place for simple approaches that can be more accessible at diverse clinical interfaces. While stratifying patients with diabetes into discrete subtypes might result in information loss compared to continuous risk modelling^39^, discrete clusters might facilitate informing clinical decisions^41^.

### Limitations

The limitations of this review reflect the limitations of the literature. To manage the breadth of literature analysed in this systematic review, we focussed on studies incorporating ‘genomic data’ rather than proteomics or metabolomics. We also did not include studies in participants at risk of type 2 diabetes, although we recognise that a body of evidence is emerging to stratify type 2 diabetes incidence risk using multiple approaches that are similar to those for established type 2 diabetes as reported here. Since we focused on studies that attempted to subgroup type 2 diabetes, we also did not capture analyses of independent cohorts with a particular type 2 diabetes phenotype at baseline, for example, studies of young people with type 2 diabetes or those with ketosis-prone type 2 diabetes (as outlined above).

### Next Steps and Recommendations

Future research should aim to identify and validate clinically useful and cost-effective methods for type 2 diabetes subclassification that can be applied across diverse populations. Such research will involve replication of a given approach in independent datasets, including from diverse ancestral populations, to ensure generalisability. For simple stratification approaches there is still much that can be done - agreement on standardised study designs for precision diagnostics studies could be a first step.

Prospective studies focusing on stratification of new-onset diabetes cases may provide valuable insights into the progression and development of distinct subtypes. Once a given approach has been appropriately replicated, additional research is required to demonstrate clinical benefit gained by applying a given subclassification approach, ideally in the form of clinical trials, and testing in diverse populations

Finally, as ML approaches require real-time computation, the development of strategies to overcome resource constraints in implementing these methods should be explored.

## Conclusion

In this first systematic review of the evidence underpinning type 2 diabetes subclassification, multiple approaches have been identified under the banner of simple criteria based on fundamental categorisation of mostly routine measures and complex approaches with multitrait (including genetic) inputs that require ML or computation. Whilst simple approaches are likely to be more easily deployed in the future, the study designs and level of evidence currently limits any firm conclusions regarding the utility of such approaches. The clinical variables and data incorporated into ‘complex’ approaches have yielded reproducible subclassifications and a growing body of evidence supports clinically meaningful associations to outcomes and treatment responses. This is a rapidly evolving field with higher quality evidence emerging all the time. It is clear that clinically relevant heterogeneity in type 2 diabetes presentations and progression is ripe for precision medicine interventions. It will be crucial to develop interventions that target diverse populations and that shall be feasible in all resource settings to prevent widening existing inequalities in the care of people with type 2 diabetes.

## Author contributions

Review Design: ALL

Systematic Review Implementation: ALL

Full-text data extraction’: ALL

Data synthesis: SM, RW, BO, JBM, MU, MS, KP, CW, MSG, JCF, RK, MRR, ASW

Manuscript writing: SM, RW, JBM & MU

Manuscript Review: ALL

Project Management: SM, RW, JBM & MU

## Supporting information

All extracted papers

## Data Availability

All data produced in the present work are contained in the manuscript

## Acknowledgements

The ADA/EASD Precision Diabetes Medicine Initiative, within which this work was conducted, has received the following support: The Covidence licence was funded by Lund University (Sweden) for which technical support was provided by Maria Björklund and Krister Aronsson (Faculty of Medicine Library, Lund University, Sweden). Administrative support was provided by Lund University (Malmö, Sweden), University of Chicago (IL, USA), and the American Diabetes Association (Washington D.C., USA). The Novo Nordisk Foundation (Hellerup, Denmark) provided grant support for in-person writing group meetings (PI: L Phillipson, University of Chicago, IL).

