## Supplementary methods for "Systematic review of precision subclassification of type 2 diabetes"

**Question 1 Search Terms**

#1
"Diabetes Mellitus, Type 2"[Mesh]
=154385

#2
type 2 diabetes[Title/Abstract] OR niddm[Title/Abstract] OR t2dm[Title/Abstract] OR t2d[Title/Abstract] OR "noninsulin dependent diabetes"[Title/Abstract] OR "non insulin dependent diabetes"[Title/Abstract] OR "type 2"[Title/Abstract] OR "type ii"[Title/Abstract] OR type2[Title/Abstract] OR typeii[Title/Abstract] OR adult-onset diabetes[Title/Abstract] OR prediabetic[Title/Abstract] OR pre-diabetic[Title/Abstract] OR Non-insulin dependent diabetes mellitus
=378950

#3
#1 OR #2
=378950

#4
"Classification"[Mesh] OR "classification" [Subheading]
=753381

#5
classificat*[Title/Abstract] OR sub classificat*[Title/Abstract] OR subclassificat*[Title/Abstract] OR subgroup*[Title/Abstract] OR "sub-group*'"OR subtype*[Title/Abstract] OR "glucose response curve"[Title/Abstract] OR "body shape index"[Title/Abstract] OR "fat evaluation"[Title/Abstract] OR "fasting insulin resistance index"[Title/Abstract] OR metabolic basis[Title/Abstract] OR immunologic basis[Title/Abstract] OR immunological basis[Title/Abstract]
=850738

#6
#4 OR #5
=1494817

#7
((("Precision Medicine"[Mesh]) OR "Genetic Heterogeneity"[Mesh]) OR "Phenotype"[Mesh]) OR "Blood Glucose"[Mesh]
=535079

#8
"precision medicine"[Title/Abstract] OR heterogeneity[Title/Abstract] OR variability[Title/Abstract] OR phenotyp*[Title/Abstract] OR pattern*[Title/Abstract] OR severity[Title/Abstract] OR feature*[Title/Abstract] OR characteristic*[Title/Abstract] OR predict*[Title/Abstract] OR stage*[Title/Abstract] OR glycemia[Title/Abstract] OR glycaemia[Title/Abstract] OR "perzonalised medicine"[Title/Abstract] OR "personalised medicine"[Title/Abstract] OR forecast*[Title/Abstract] OR determinant*[Title/Abstract]
=7227358

#9
#7 OR #8
=7484521

#10
"Medical History Taking"[Mesh]
=22396

#11
family history[Title/Abstract] OR relative[Title/Abstract]
=993615

#12
#10 OR #11
=1013777

#13
#9 OR #12
=8116688

#14

((((((((((((((("Insulin-Secreting Cells"[Mesh]) OR "Islets of Langerhans"[Mesh]) OR "C-Peptide"[Mesh]) OR "Glucose Tolerance Test"[Mesh]) OR "Glucose Clamp Technique"[Mesh]) OR "Biomarkers"[Mesh]) OR ( "Body Mass Index"[Mesh] OR "Obesity"[Mesh] OR "Overweight"[Mesh] )) OR "Insulin Resistance"[Mesh]) OR "Body Weight"[Mesh]) OR "Glycemic Index"[Mesh]) OR "Metabolic Syndrome"[Mesh]) OR "Lipids"[Mesh]) OR "Cholesterol"[Mesh]) OR "Dyslipidemias"[Mesh]) OR "Body Fat Distribution"[Mesh]) OR "Intra-Abdominal Fat"[Mesh]
=2638388

#15
Beta cell*[Title/Abstract] OR betacell*[Title/Abstract] OR beta-cell*[Title/Abstract] OR beta cell dysfunction[Title/Abstract] OR beta cell function[Title/Abstract] OR insulin secretion[Title/Abstract] OR c-peptide[Title/Abstract] OR "first phase"[Title/Abstract] OR "glucose tolerance test"[Title/Abstract] OR ogtt[Title/Abstract] OR ivgtt[Title/Abstract] OR clamp[Title/Abstract] OR biomarker*[Title/Abstract] OR biological marker*[Title/Abstract] OR "body mass index"[Title/Abstract] OR bmi[Title/Abstract] OR over weight[Title/Abstract] OR overweight[Title/Abstract] OR obesity[Title/Abstract] OR adiposity[Title/Abstract] OR "insulin resistance"[Title/Abstract] OR homa[Title/Abstract] OR homeostasis model assessment[Title/Abstract] OR weight[Title/Abstract] OR score[Title/Abstract] OR index[Title/Abstract] OR metabolic syndrome[Title/Abstract] OR excess fat mass[Title/Abstract] OR lipid*[Title/Abstract] OR triglycerid*[Title/Abstract] OR cholesterol[Title/Abstract] OR dyslipidemia[Title/Abstract] OR body fat distribution[Title/Abstract] OR bodyfat distribution[Title/Abstract] OR ketosis[Title/Abstract] OR atypical[Title/Abstract] OR "age at onset"[Title/Abstract] OR "age at diagnosis"[Title/Abstract] OR ketoacidosis[Title/Abstract]
=3645559

#16
#14 OR #15
=5129710

#17
#3 AND #6 AND #13 AND #17
=7208

#18
#17 Filters:Humans
=5993

#19
(review[Publication Type] OR "systematic review"[Publication Type] OR (editorial[Title/Abstract] OR review[Title/Abstract])
=2902309

#20
#18 NOT #19
=**4825 references**
 **Question 2 Search Terms**

#1
Diabetes mellitus/classification[MeSH terms] OR diabetes mellitus type 2/classification[Mesh Terms] OR diabetes mellitus type 2/epidemiology[MeSH Terms] OR diabetes mellitus type 2/physiopathology[MeSH Terms] OR diabetes mellitus type 2[MeSH Terms] OR diabetes complications[MeSH Terms] OR Prediabetes[MeSH Terms]
=280913

#2
"Diabetes mellitus, type 2"[Title/Abstract] OR T2D[Title/Abstract] OR "diabetes mellitus type-2" prediabetes[Title/Abstract] OR pre-diabetes IGT[Title/Abstract] OR IFC[Title/Abstract] OR type 2 diabetes[Title/Abstract] OR cluster based diabetes[Title/Abstract] OR cluster-based diabetes[Title/Abstract] OR "adult onset diabetes"[Title/Abstract] OR adult-onset diabetes[Title/Abstract] Filters: Humans
=116423

#3
"adult-onset diabetes subgroups"
=5716

#4
#1 OR #2 OR #3
=302899

#5
((((("Phenotype"[Mesh]) OR "Body Mass Index"[Mesh]) OR "Autoantibodies"[Mesh]) OR "Blood Glucose"[Mesh]) OR "Glycated Hemoglobin A"[Mesh]) OR "Hypoglycemic Agents"[Mesh]
=809305

#6
(((((("Genetic Loci"[Mesh]) OR "Genetic Markers"[Mesh]) OR "Genetic Predisposition to Disease"[Mesh]) OR "Genome-Wide Association Study"[Mesh]) OR "Bayes Theorem"[Mesh]) OR "Founder Effect"[Mesh]) OR "Multigene Family"[Mesh]
=366199

#7
classification[Title/Abstract] OR phenotyp*[Title/Abstract] OR subphenotyp*[Title/Abstract] OR subtype*[Title/Abstract] OR "clinical type*"[Title/Abstract] OR autoantibod*[Title/Abstract] OR "blood glucose"[Title/Abstract] OR "glycated hemoglobin A"[Title/Abstract] OR "genetic loci"[Title/Abstract] OR "genetic predisposition to disease"[Title/Abstract] OR "genome-wide association study"[Title/Abstract] OR "founder effect*"[Title/Abstract] OR "bayes theorem"[Title/Abstract] OR HOMA*[Title/Abstract] OR subgroup*[Title/Abstract] OR HbA1c[Title/Abstract] OR A1C[Title/Abstract]
=1681470

#8
#5 OR #6 OR #7
=2391509

#9
((("Cluster Analysis"[Mesh]) OR ( "Supervised Machine Learning"[Mesh] OR "Unsupervised Machine Learning"[Mesh] OR "Machine Learning"[Mesh] )) OR "Artificial Intelligence"[Mesh]) OR "Monitoring, Physiologic"[Mesh]
=398710

#10
cluster analysis[Title/Abstract] OR machine learning[Title/Abstract] OR reinforcement learning[Title/Abstract] OR deep learning[Title/Abstract] OR CNN[Title/Abstract] OR "convolutional neural network"[Title/Abstract] OR "data driven cluster*"[Title/Abstract] OR "data-driven cluster*"[Title/Abstract] OR "physiologic monitor*"[Title/Abstract] OR "artificial intelligence"[Title/Abstract] OR AI[Title/Abstract] OR clusters[Title/Abstract] OR subgroup*[Title] OR subtype*[Title]
=365087

#11
#9 OR #10
=689538

#12
#4 AND #8 AND #11
=5986
